# Cervicovaginal microbiota predicts *Neisseria gonorrhoeae* clinical presentation

**DOI:** 10.1101/2021.10.07.21264698

**Authors:** Angela Lovett, Arlene C. Seña, Andrew N. Macintyre, Gregory D. Sempowski, Joseph A. Duncan, Andreea Waltmann

## Abstract

*Neisseria gonorrhoeae* infection of the female lower genital tract can present with a spectrum of phenotypes ranging from asymptomatic carriage to symptomatic cervical inflammation, or cervicitis. The factors that contribute to the development of asymptomatic or symptomatic infections are largely uncharacterized. We conducted a pilot study to assess differences in the cervicovaginal microbial community of patients presenting with symptomatic *vs*. asymptomatic *N. gonorrhoeae* infections to a sexually transmitted infections (STI) clinic. DNA was isolated from cervicovaginal swab specimens from women who tested positive for *N. gonorrhoeae* infection using a clinical diagnostic nucleic acid amplification test. We performed deep sequencing of 16S ribosomal RNA gene amplicons, followed by microbiome analyses with QIIME, and species-specific real-time PCR to assess the composition of microbial communities cohabitating the lower genital tract with the infecting *N. gonorrhoeae*. Specimens collected from asymptomatic individuals with *N. gonorrhoeae* infection and no co-infection with *Chlamydia trachomatis* and/or *Trichomonas vaginalis* carried *Lactobacillus-*dominant microbial communities more frequently than symptomatic patients without co-infection. When compared to asymptomatic individuals, symptomatic women had microbial communities characterized by more diverse and heterogenous bacterial taxa, typically associated with bacterial vaginosis (BV) (*Prevotella, Sneathia, Mycoplasma hominis* and Bacterial Vaginosis-Associated Bacterium-1 (BVAB1)/”*Candidatus Lachnocurva vaginae)*. Both symptomatic and asymptomatic *N. gonorrhoeae* patients with additional STI co-infection displayed a BV-like microbial community. We used a murine model of *N. gonorrhoeae* infection in mice pre-colonized with *Lactobacillus crispatus* to test whether pre-existing *L. crispatus* was protective from *N. gonorrhoeae* colonization or whether *N. gonorrhoeae* infection could drive the loss of *L. crispatus* during infection. Vaginal infection with either *N. gonorrhoeae* strain 1291 or an isogenic mutant known to exhibit lower inflammatory had no impact on *Lactobacillus* burden recovered from the mice. These data taken together suggest that *Lactobacillus-*dominant vaginal microbial community may protect individuals from developing symptoms during lower genital tract infection with *N. gonorrhoeae*.

## Introduction

*Neisseria gonorrhoeae* is a sexually transmitted bacterial pathogen responsible for 90 million infections globally each year [1]. *N. gonorrhoeae* is a strictly human pathogen and infections are typically localized to the lower genital tract. Although acute symptomatic infection is often recognized and treated with antibiotic therapy, a surprisingly large proportion of *N. gonorrhoeae* infections are asymptomatic [2-4]. Upwards of 50% of lower genital tract infections in females are asymptomatic [5, 6]. While the prevailing dogma is that male urethral infections are symptomatic, compelling reports have documented that asymptomatic genital gonorrhea is prevalent in both biological sexes and across a wide range of settings [2, 7-11]. Asymptomatic infection is not without consequence. Untreated asymptomatic infections can ascend to the upper genital tract leading to health complications including pelvic inflammatory disease and infertility in women [12-14]. Asymptomatic genital *N. gonorrhoeae* poses a risk of onward transmission to sex partners and ascending infection. Mathematical models of gonorrhea transmission have confirmed unequivocally the significant contribution of subclinical infections in maintaining community transmission [15]. The determinants and mechanisms that underlie asymptomatic and symptomatic gonorrhea infection are unknown. This knowledge gap greatly hinders efforts to develop new strategies for gonorrhea control.

Symptomatic *N. gonorrhoeae* infection most commonly leads to localized host inflammation at the site of infection, urethritis in males, and cervicitis in females. *N. gonorrhoeae* itself is resistant to many host antimicrobial responses which may contribute to its ability to cause infection under these conditions of localized inflammation. During an infection, *N. gonorrhoeae* must compete with the natural microbial community at the mucosal surface to establish infection [16]. Cervicovaginal microbial communities play an important role in sexual and reproductive outcomes, including protection from pathogens, as the composition of the cervicovaginal microbiota has been shown to modify susceptibility to several sexually-transmitted pathogens (STI) [17-23]. For example, human vaginal microbiota dominated by *Lactobacillus crispatus* is associated with reduced risk of acquisition of STI, like HIV [24-27]. In addition, women with clinically apparent bacterial vaginosis (BV), a clinical condition characterized by depleted levels of of *Lactobacillus* species and an increased abundance of diverse groups of facultative anaerobes, have an increased risk of acquiring and transmitting STI, including *N. gonorrhoeae* [24, 28-30] and at an increased risk of adverse reproductive and obstetric outcomes [31-33], irrespective of whether the BV is symptomatic or not [17, 33, 34]. These factors indicate a mechanistic contribution of *L. crispatus* to protection from STI, presumably through the production of lactic acid and thus the maintenance of a low-pH vaginal microenvironment [35]. The reasons why the microbiota of some women is dominated by *Lactobacillus* species, whereas that of others becomes dominated by anaerobes are not completely understood.

Molecular definitions of the cervicovaginal microbiota of reproductive age women, with respect to microbial composition and diversity, are now standard [21, 27, 36]. Using 16S rRNA sequencing or metagenomics, distinct cervicovaginal microbial community types (CTs) have been classified [21, 27, 36]. The “optimal types” CT1 and CT2 are dominated by *L. crispatus* and *L. gasseri*, respectively. The other two *Lactobacillus*-dominant molecular community types are *L. iners* (CT3) and *L. jensenii* (CT5). Notably, not all *Lactobacillus*-dominant types are equal in their protective effect against STI, as *L. iners*-dominated vaginal microbiotas may actually place patients at higher risk of STI infection, like chlamydia or HIV, when compared to *L. crispatus*-dominant microbiotas [25-27]. Finally, CT4 communities are characterized by a diverse and heterogenous group of anaerobes (e.g. *Atopobium, Prevotella, Dialister, Gardnerella, Megasphaera, Peptoniphilus, Sneathia, Eggerthella, Aerococcus, Finegoldia*, and *Mobiluncus* [36]) and this vaginal environment is a significant risk factor for having clinically-diagnosed bacterial vaginosis (BV) [37-39] and is termed molecular-BV [34].

Studies examining the impact of vaginal microbiomes on inflammatory states found that women with clinical-BV or molecular BV (i.e. CT4 microbial communities) had higher cervicovaginal levels of pro-inflammatory cytokines (IL-1α, IL-1β, IL-6, IL-12 and IL-8) than BV-negative or *Lactobacillus*-dominant women, respectively [22, 27, 40]. Notably, the more the vaginal microbiota shifts away from a state dominated by *L. crispatus* towards dysbiosis (i.e. towards CT3 and CT4 types) the more marked the inflammation [21, 41, 42], independently of concurrent STIs, including gonorrhea [21]. This indicates that the variations in vaginal microbial diversity that are common in women with BV could influence inflammatory responses that characterize symptomatic *N. gonorrhoeae* infection.

We sought to understand whether differences in microbial composition of the genital tract were associated with symptomatic or asymptomatic presentation of *N. gonorrhoeae* infection, by utilizing 16S ribosomal RNA (rRNA) amplicon deep sequencing of clinician-collected cervicovaginal specimens from females diagnosed with gonorrhea. Because the causal relationship between gonorrhea infection, development of symptoms, and presence of *L. crispatus* remains an open question in the field, we conducted experiments with controlled infection models of *N. gonorrhoeae* in mice pre-colonized to test whether *N. gonorrhoeae* infection could drive the loss of pre-existing *L. crispatus* during infection or if .*L. crispatus* differentially protected against infection with *N. gonorrhoeae* strains with different inflammatory potential.

## Methods

### Study Population and Sample Collection

Specimens used in this study were remnant specimens collected from female patients that attended a public sexually transmitted infection (STI) clinic located in Durham, North Carolina in 2011 and tested positive for *Neisseria gonorrhoeae* using the clinical diagnostic assay Aptima Combo 2® assay (for CT/NG) by Hologic. Clinician-collected cervicovaginal swabs used for diagnosis were collected as per routine care prior to any treatment. The age, race, reported symptoms, and diagnosis for each study subject were linked to each specimen by a study clinician when a positive specimen was identified. The de-identified remnant cervical swab samples were stored in Aptima buffer in their respective transport tubes at −80°C until DNA extraction was performed.

### DNA Extraction

200 μL of Aptima buffer from each sample were transferred to sterile 2 mL tubes containing 200 mg of ≤100 μm glass beads (Sigma), 0.3 mL of 20mg/mL lysozyme solution (Thermo Fisher) and 0.3 mlL of Qiagen ATL buffer. Bead-beating was then carried out for 10 minutes in a Qiagen TissueLyser II at 30Hz to ensure optimal DNA yield from Gram-positive bacteria. Subsequently, samples were incubated at 37°C for 30 minutes. After a brief centrifugation, supernatants were aspirated and transferred to a new sterile tube with Qiagen AL buffer containing Proteinase K (600 IU/μL). Samples were then incubated at 70°C for 10 minutes. DNA was purified using a standard on-column purification method using Zymo-spin mini columns and Qiagen buffers AW1 and AW2 as washing agents. DNA was eluted in 100 μL of 10 mM Tris (pH 8.0).

### 16S Ribosomal RNA Sequencing

For amplicon library preparation we used fusion primers composed of Ion Torrent adapter 5’-*CCATCTCATCCCTGCGTGTCTCCGACTCAG* -3’ for the forward primer and 5’-*CCTCTCTATGGGCAGTCGGTGAT* -3’ for the reverse primer, and universal bacterial primer 8F 5’-*AGAGTTTGATCCTGGCTCAG*-3’ and 338R 5’-*GCTGCCTCCCGTAGGAGT*-3’. The forward primer also included a 10 bp IonXpress™ barcode, unique to each sample. Each bacterial DNA sample was run in duplicate in a 25 μL PCR reaction containing: 4 μL of 5x MyTaq Reaction Buffer (Bioline); 0.6 μL each of 15 μM Forward Primer and 15 μM Reverse Primer (Integrated DNA Technologies); 0.5 μL MyTaq HS DNA Polymerase (Bioline); 100 ng template DNA; water to 25 μL. Samples were denatured at 94°C for 5 minutes, followed by 35 cycles of 94°C for 45 seconds, 55°C for 45 seconds and 72°C for 90, followed by an extension at 72°C for 10 minutes and a 4°C hold. Each sample was visualized on a 2 % agarose gel. Bands were excised and duplicate bands were combined into one tube. Gel purification was performed using the Qiagen Gel Extraction Kit (Qiagen) according to the manufacturer’s protocol. Samples were quantified using an Agilent 2100 Bioanalyzer (Agilent). Quantification information was used to create a library by combining equimolar concentrations of each sample. The prepared library was sequenced on the Ion Torrent PGM Instrument (Life Technologies) according to the manufacturer’s protocol at UNC-CH High Throughput Sequencing Facility.

### Sequence Data Analysis

Sequencing output was demultiplexed and the resulting paired-end reads were joined using the QIIME 1.9.0 [43] by invocation of fastq-join with the default parameters. Index and linker primer sequences were trimmed, and the reads were subsequently filtered for quality using a sliding window of 50 bases, moving by 5 bases, requiring an average quality score of 20 or above. Quality control of both raw and processed sequencing reads was verified by FastQC [44]. Sequences were clustered into operational taxonomic units (OTU) based on the *de novo* OTU picking algorithm using the QIIME implementation of UCLUST [45] at a similarity threshold of 97%. OTUs identified as chimeric by vsearch [46] of the ChimeraSlayer “gold” reference database [47] and those composed of a single read (singletons) were eliminated. The remaining OTUs were assigned taxonomic identifiers with respect to the Greengenes database [48], their sequences were aligned using template alignment through PyNAST [49], and a phylogenetic tree was built with FastTree 2.1.3 [50]. If after the Greengenes taxonomic assignment a taxon of interest was ambiguous at the genus level or when putative species taxonomy was sought, we consulted the 16S sequences in the National Center for Biotechnology Information (NCBI) Genbank repository with the Basic Local Alignment Search Tool (BLAST) [51] and/or with the multiple sequence comparison by log-expectation method (MUSCLE) implemented in Geneious using reference genomes.

### Microbiome Analyses

Alpha diversity was measured by three different metrics (Chao1; and observed species; phylogenetic diversity, PD) using QIIME. Beta diversity estimates were calculated within QIIME using weighted and unweighted Unifrac distances [52] between samples. Results were summarized and visualized through principal coordinate analysis (PCoA) in QIIME. Cervicovaginal community types (CTs) were assigned using established definitions [21, 27, 36] based on diversity and relative abundance of bacterial taxa. These definitions classify samples with relative majority abundance assigned to *Lactobacillus crispatus, L. gasseri, L. iners* or *L. jensenii* as CT1, CT2, CT3, CT5, respectively. According to the same definitions, low *Lactobacillus* communities comprising a diverse and heterogenous group of anaerobes (e.g. *Atopobium, Prevotella, Dialister, Gardnerella, Megasphaera, Peptoniphilus, Sneathia, Eggerthella, Aerococcus, Finegoldia, and Mobiluncus* are classified as CT4 or “molecular bacterial vaginosis” [34].

### Microbial DNA qPCR Array

The Vaginal Flora Microbial DNA qPCR Array (Cat. no. 330261 BAID-1902Y, Qiagen) was used for vaginal microbiome profiling. Each specimen was processed and run on the 96 well array plate format. The array contained assays for the detection of 90 microbial species, 2 Pan bacteria controls (Pan Bacteria 1, Pan Bacteria 3), 1 Pan Fungi control (Pan *Aspergillus/Candida*), 2 host controls (Hs/Mm.GAPDH Hs/Mm.HBB1) and a positive PCR control (PPC). 500 ng of genomic DNA was mixed with 1,275 μL microbial qPCR Mastermix and water as needed to bring the total volume to 2,550 μL, using the manufacturer’s instructions. Individual reaction mix aliquots of 25 μL were added to each well of the plate, the array plate was tightly sealed, centrifuged at 1000 rpm for 1 min and loaded onto the Real-Time PCR machine. PCR was performed with an initial PCR activation step at 95°C for 10 minutes, followed by 2-step cycling of denaturation for 15 seconds at 95°C, with annealing and extension for 2 minutes at 60 °C for 40 cycles. The CT values for each well were imported into the Microbial DNA qPCR Array Excel template (Qiagen) for analysis. The ΔCT between the patient sample DNA and no DNA template negative control was calculated for each set of species-specific primer sets, following manufacturer’s instructions for analysis. A species was reported as “present, +” if ΔCT was >3, an 8-fold increase in signal over background.

### Murine Infection Models

The protocol for murine vaginal *N. gonorrhoeae* infection was adapted from a model previously described by Jerse *et al*. [53]. In brief, 4 to 6 week-old female BALB/c mice were synced in the proestrous stage of their estrous cycle by administering β-estradiol on days -2, 0 and 2 of infection. Mice were also given daily intraperitoneal injections of streptomycin sulfate to promote infection through reduction of commensal organisms. *L. crispatus* was acquired from the American Tissue Culture Collection (ATCC, 33820™) and was used to establish the pre-colonized *Lactobacillus* vaginal model. Mouse infection with *N. gonorrhoeae* strains 1291 (wild-type) and 1291Δ*msbB* (mutant) have been previously described [54]. Prior to infection, bacterial strains were grown overnight (18 hour) on De Man, Rogosa and Sharpe (MRS) agar for *L. crispatus* or Gonococcal Base (GCB) agar for *N. gonorrhoeae* at 37°C in 5% CO _2_, harvested and transferred into PBS for inoculation. Bacterial suspensions were quantified by measuring the optical density at 600 nm (OD_600_). Mice were pre-colonized with *L. crispatus* by vaginal inoculation on day -1 with an estimated 10^8^ CFUs of *L. crispatus* prepared in PBS. Mice were subsequently infected with *N. gonorrhoeae* (1291 or 1291-Δ*msbB)* by vaginal inoculation with an estimated 10^6^ CFUs of the indicated N. gonorrhoeae strain in PBS on day 0. Vaginal swabs were collected daily from each mouse for 10 days following inoculation with *N. gonorrhoeae*. The quantity of *L. crispatus* was determined by plating of serial dilutions of the vaginal swabs on MRS agar. The quantity *N. gonorrhoeae* was determined by plating of serial dilutions of the vaginal swabs on (GCB) agar supplemented with vancomycin, colistin, nystatin, trimethoprim (VCNT).

### Statistical Analyses

Statistical analyses were performed in PRISM 9 or STATA 16. Differences in donor characteristics and between sample groups were investigated using the Fisher’s exact test. We compared α-diversity between sample groups with non-parametric two-sample t*-*tests using 1000 Monte Carlo permutations to calculate the p*-*values. To test whether sample groups were statistically different we used non-parametric ANOSIM (ANalysis Of Similarities) tests and non-parametric two-sample *t*-tests with 1000 Monte Carlo permutations to derive p-values implemented in QIIME. Kaplan-Meier curves were used to estimate clearance of *N. gonorrhoeae* and time to clearance (in days) in the mouse model of infection. For comparisons between multiple groups one way ANOVA was used, correcting for multiple comparisons with Tukey post hoc tests.

## Results

### Cohort characteristics of *N. gonorrhoeae* infected women

In this pilot study, we used remnant nucleic acid material from cervical swabs collected from a convenience sample of 19 women deemed to be *N. gonorrhoeae-*positive by Aptima clinical diagnostic testing. Of these, ten individuals reported symptoms to the provider (defined as symptomatic) and nine did not (defined as asymptomatic) (Table 1). The reported symptoms at the time of cervical swab sampling included vaginal discharge (9/10, 90.0%), genital irritation (1/10, 10.0%), and dysuria (2/10, 20%). There was a trend for younger women to report symptoms (p=0.090). Of the 19 specimens, 17 (89.5%) were collected from women who identified as having African American race. Among the African American women, the proportion of symptomatic and asymptomatic individuals was comparable (p=0.211). The presence or absence of *C. trachomatis*, another STI pathogen, was assessed by NAAT testing in conjunction with clinical *N. gonorrhoeae* testing, and no difference in *C. trachomatis* prevalence was observed between asymptomatic and symptomatic presentation (p=0.590).

**Table 1.** Study population characteristics at baseline. The age, race, and self-reported symptoms of 19 women positive for *N. gonorrhoeae* infection by clinical test who presented at a local STI clinic are provided.

| Baseline characteristics | Symptomatic | Asymptomatic | P-value |
| --- | --- | --- | --- |
|  | n = 10 | n = 9 |  |
| Age, mean (range) | 21.1 (15-37) | 26.6 (16-40) | 0.09 |
| Race (n) | Black (10) | Black (7) | n/a |
|  | Latina (0) | Latina (1) |  |
|  | White (0) | White (1) |  |
| Symptoms (%) | Vaginal Discharge (90.0) | None (100.0) | n/a |
|  | Genital irritation (10.0) |  |  |
|  | Dysuria (20.0) |  |  |

Since other STI pathogens could also be responsible for causing lower genital tract symptoms (e.g. *Trichomonas vaginalis* or *Mycoplasma genitalium*), we used a commercial microbial qPCR array to test for the presence of other STI pathogens (Table 2). Specimens from two women did not provide evaluable results due to insufficiently recovered DNA material. Among the 17 specimens that gave analyzable results on this array, 6 of 7 (85.7%) asymptomatic specimens and 8 of 10 (80.0%) symptomatic specimens had detectable *Neisseria* species DNA (Table 2). Because these specimens all tested positive for *N. gonorrhoeae* using the APTIMA Combo 2, these results indicate that the sensitivity of the microbial qPCR array may be lower for detecting *N. gonorrhoeae* in this specimen type than the APTIMA Combo 2. *C. trachomatis* was detected in 3 of 3 specimens that were positive by APTIMA Combo 2 test and had evaluable qPCR results in the qPCR array (Table 2). *T. vaginalis* was detected in 4 of 10 (40.0%) evaluable symptomatic individuals and 2 of 7 (28.6%) asymptomatic individuals (Table 2). *M. genitalium* was not detected by qPCR array in any of the specimens (Table 2). When accounting for both clinical *C. trachomatis* testing by APTIMA Combo 2 and real-time PCR array results, the proportion of STI co-infection with *C. trachomatis* or *T. vaginalis* was not significantly different between symptomatic (5/10, 50.0%) and asymptomatic (5/9, 55.6%) individuals (p=0.625). Because co-infection with other STI pathogens was not associated with symptomatic presentation we next sought to assess whether the non-STI vaginal microbial community was associated with the presence or absence of symptoms.

**Table 2.**
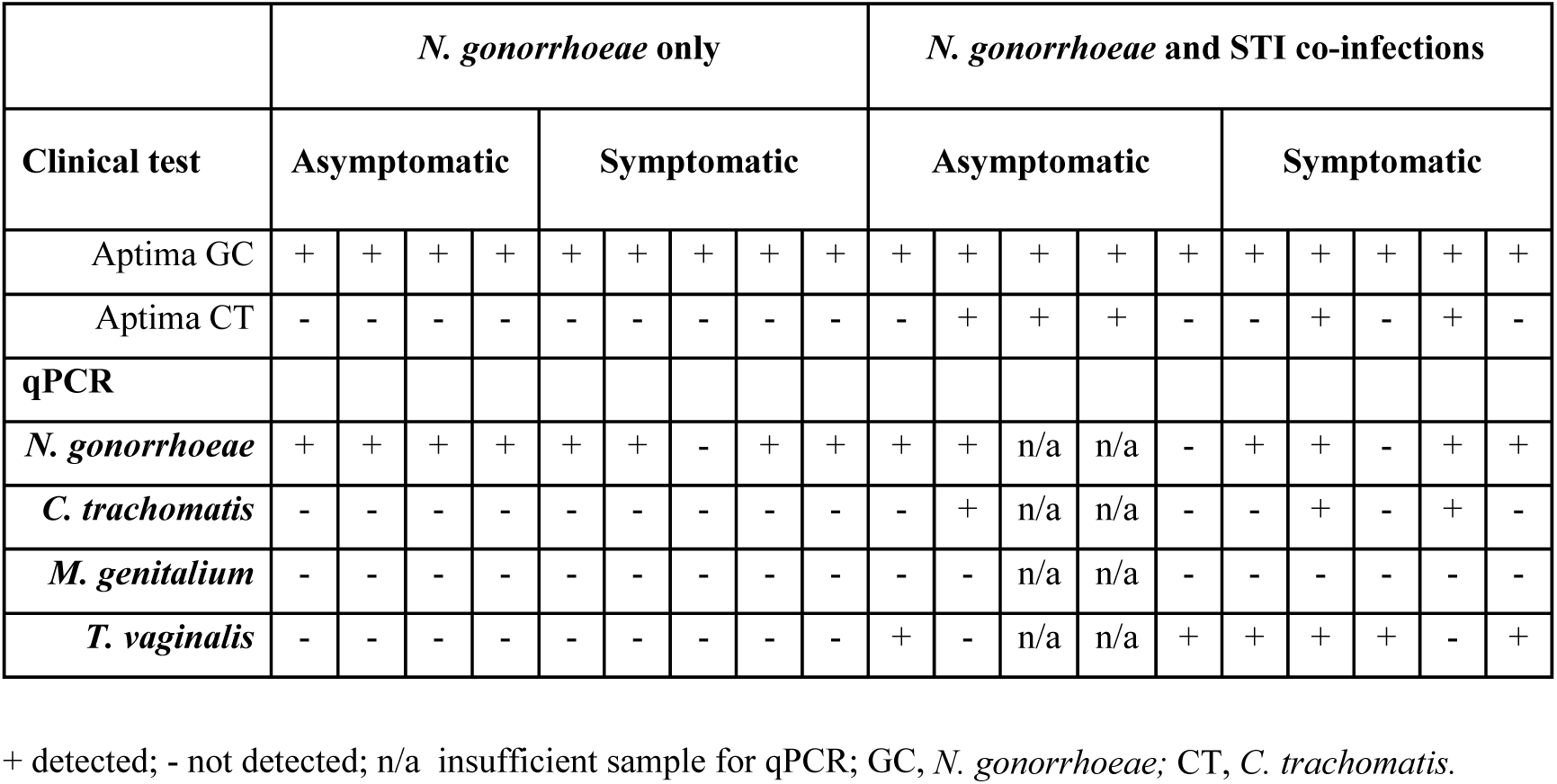
Results of clinical Aptima (NAAT) test results and microbial DNA quantitative real-time PCR (qPCR) array. Each column across the categories reflects one individual participant.

### *Neisseria spp*. abundance represents a small proportion of bacterial communities in both symptomatic and asymptomatic patients

The genital microbial community of the 19 study participants was characterized with 16S amplicon deep sequencing. A total 157,006 paired-end reads were obtained. After demultiplexing and elimination of low-quality reads, 100,216 reads were retained for downstream analyses of alpha diversity and beta diversity (mean number of reads per sample = 5,274; range = 2,470–9,528 reads). Paired reads were deposited in the Sequence Read Archive (SRA) under the accession PRJNA768436. The individual microbial communities of *N. gonorrhoeae* infected patients were compared to those who presented with and without symptoms. Because other STI pathogens might be associated with different microbial community profiles, we also compared specimens from individuals without co-infecting *C. trachomatis* or *T. vaginalis* (by clinical test and/or real-time PCR) separately from those with co-infection. We first examined whether the relative abundance of *Neisseria ssp*. assigned reads was associated with symptomatic presentation. *Neisseria*-assigned reads made up only 0.24% of all reads in the dataset and were a minor component of the bacterial community in each individual (Figure 1). In this limited set of specimens, the point estimate of the relative of abundance of reads from *Neisseria spp*. was highest in symptomatic individuals without *C. trachomatis* or *T. vaginalis* co-infection, though no significant difference in relative abundance between any group was observed (Figure 1).

**Figure 1.**
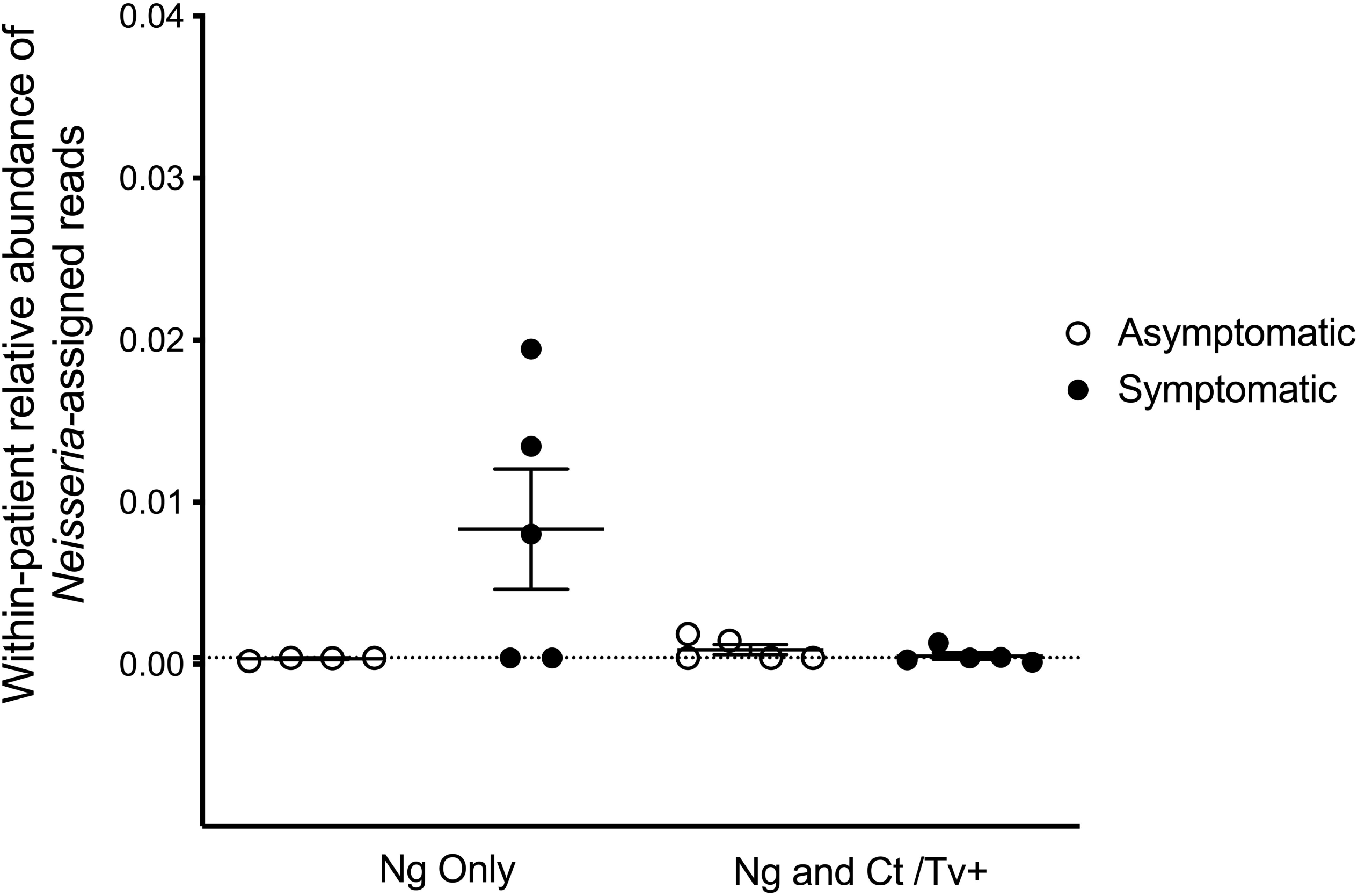
Reads assigned to the *Neisseria* genus comprise a small proportion of the cervicovaginal microbial community. We characterized the genital microbial communities of 19 females with clinically-confirmed gonorrhea using 16S amplicon deep sequencing and microbiome analysis with QIIME. After demultiplexing and eliminating low-quality reads, 100,216 reads were retained for downstream analyses (mean number of reads per sample = 5,274; range = 2,470–9,528 reads). *Neisseria*-assigned 16S ribosomal DNA reads made up only 0.24% of all reads in the dataset and are plotted for each individual’s microbial community. No significant differences in relative abundance of 16S reads from *Neisseria spp* between any group was observed using one way ANOVA with Tukey’s correction for multiple comparisons.

### Microbial community diversity is different between symptomatic and asymptomatic *N. gonorrhoeae* infection

The overall alpha diversity did not differ when observed taxa, Chao1 and phylogenetic diversity were compared between individuals with symptomatic and asymptomatic *N. gonorrhoeae* infection (Figure 2A, Figure S1 A-C) and between individuals with and without other STI (Figure 2B, Figure S1 D-E). However, the number of dominant taxa comprising the majority of the microbial community (i.e., 90% of all detected taxa) was significantly lower in individuals with asymptomatic *N. gonorrhoeae* infection without STI co-infection *vs*. both individuals with symptomatic *N. gonorrhoeae* and with STI co-infection (Figure 2C).

**Figure 2.**
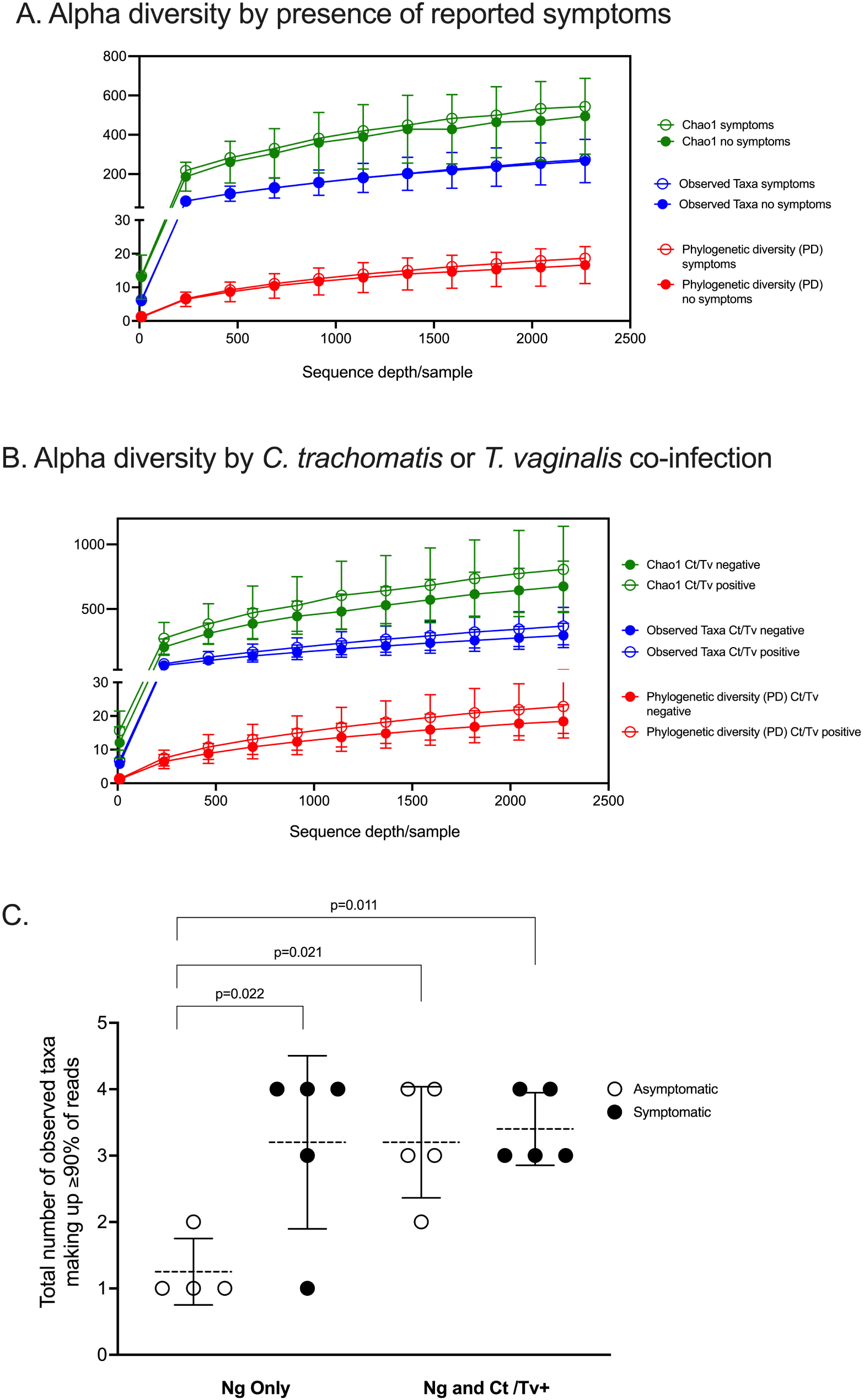
Alpha diversity analyses with respect to clinical presentation and other STI co-infection. Alpha diversity was measured by three different metrics (Chao1; and observed species; phylogenetic diversity, PD) at a depth of 2,270 reads and visualized as rarefaction plots. The overall alpha diversity including observed taxa, Chao1 and phylogenetic diversity are plotted for individuals with symptomatic and asymptomatic *N. gonorrhoeae* infection **(A)** and for individuals with and without other STI **(B)**. The number of dominant taxa comprising the majority of the microbial community (i.e., 90% of all detected taxa) is plotted for individuals with asymptomatic and symptomatic *N. gonorrhoeae* infection with and without STI co-infection **(C)**. Statistical significant differences between patient groups were explored with one way ANOVA with Tukey’s correction for multiple comparisons.

Differences between symptomatic and asymptomatic patients, but not between patients with and without *C. trachomatis* and/or *T. vaginalis* co-infection, were reflected in beta diversity analyses, with statistically significant ANOSIM tests and clear separation on PCoA plots by two different methods: weighted Unifrac ANOSIM R = 0.20, p-value = 0.032, Figure 3A) and unweighted Unifrac ANOSIM R = 0.24 p-value = 0.011, Figure 3B).

**Figure 3.**
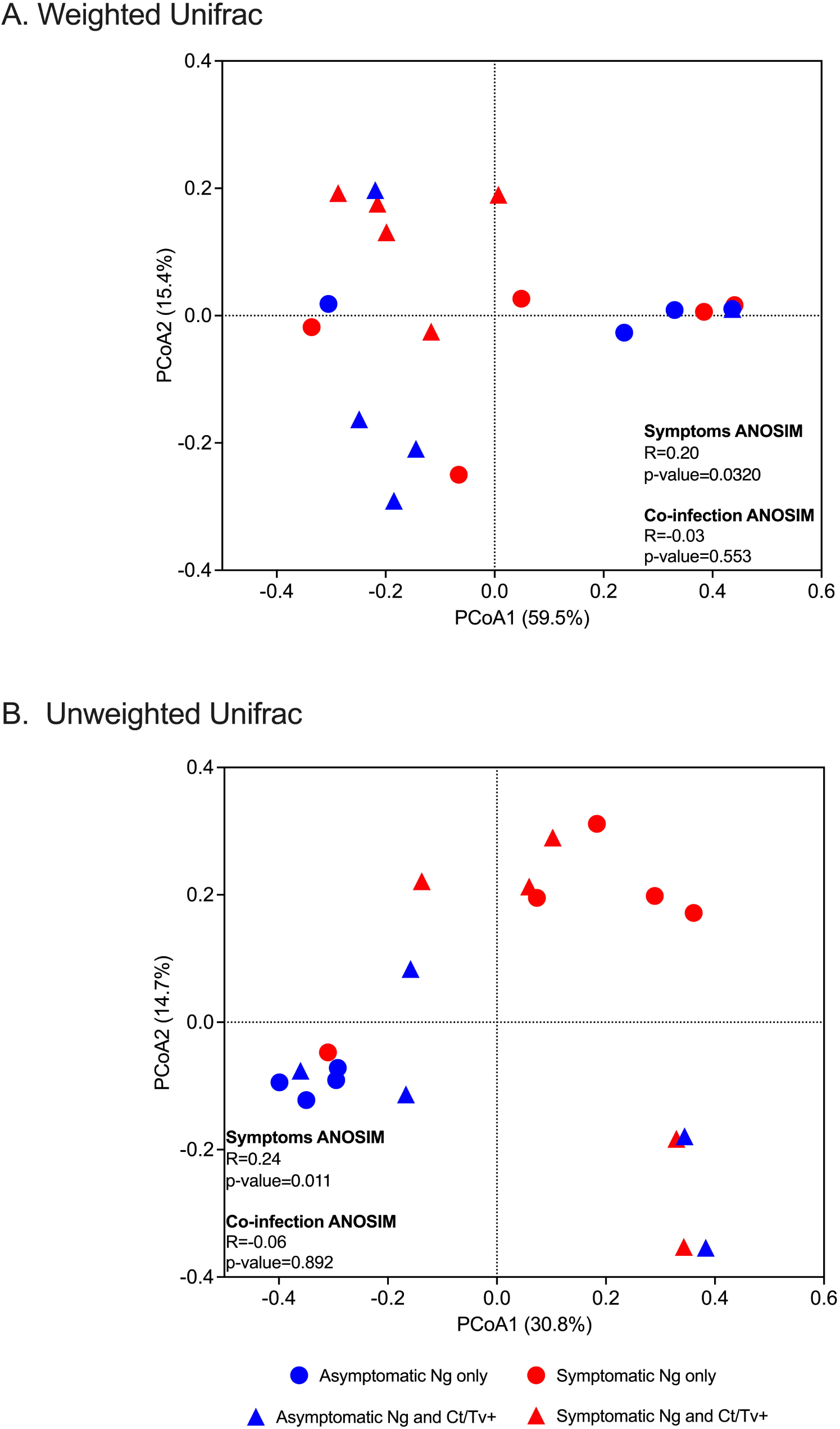
Beta diversity analyses with respect to clinical presentation and other STI co-infection. Beta diversity estimates were calculated within QIIME using weighted **(A)** and unweighted Unifrac **(B)** distances between samples. Results were summarized and visualized through principal coordinate analysis (PCoA) in QIIME. ANOSIM tests were used to assess the strength of the clustering patterns and statistical significance. Differences between symptomatic and asymptomatic patients, but not between patients with and without *C. trachomatis* and/or *T. vaginalis* co-infection, were found, with clear separation on PCoA plots by two different methods: weighted Unifrac ANOSIM R = 0.20, p-value = 0.032, Figure 3A) and unweighted Unifrac ANOSIM R = 0.24 p-value = 0.011).

### Asymptomatic patients without an STI co-infection are more frequently characterized by low diversity, *Lactobacillus*-dominant genital communities

The relative abundance of *Lactobacillus spp*. assigned reads differed by patient group. The distribution of *Lactobacillus spp*. relative abundances was: 92.2% among asymptomatic individuals with no co-infection, 35.3% among asymptomatic individuals with co-infection, 21.6% among symptomatic individuals with no co-infection, and 11.5% among symptomatic individuals with co-infection (Figure 4A). This was evident when inspecting the individual taxa plots (Figure 4B), as the four asymptomatic patients with only *N. gonorrhoeae* infection and no detected co-infection were dominated by *Lactobacillus* taxa, whereas *Lactobacillus*-predominance was observed less frequently in specimens from women with symptomatic *N. gonorrhoeae* infection regardless of the presence of additional STI (2/10, 20.0% symptomatics *vs*. 6/9, 66.7% asymptomatics, p=0.040, Figure 4A). Differences in within-patient relative abundance of *Lactobacillus spp*. in symptomatics with gonorrhea only *vs*. asymptomatics with gonorrhea were statistically significant (p=0.019, Figure 5A). Similarly, the within-patient relative abundance of *Lactobacillus spp*. in symptomatics with gonorrhea only vs. that of symptomatic with co-infections also varied significantly (p=0.007, Figure 5A).

**Figure 4.**
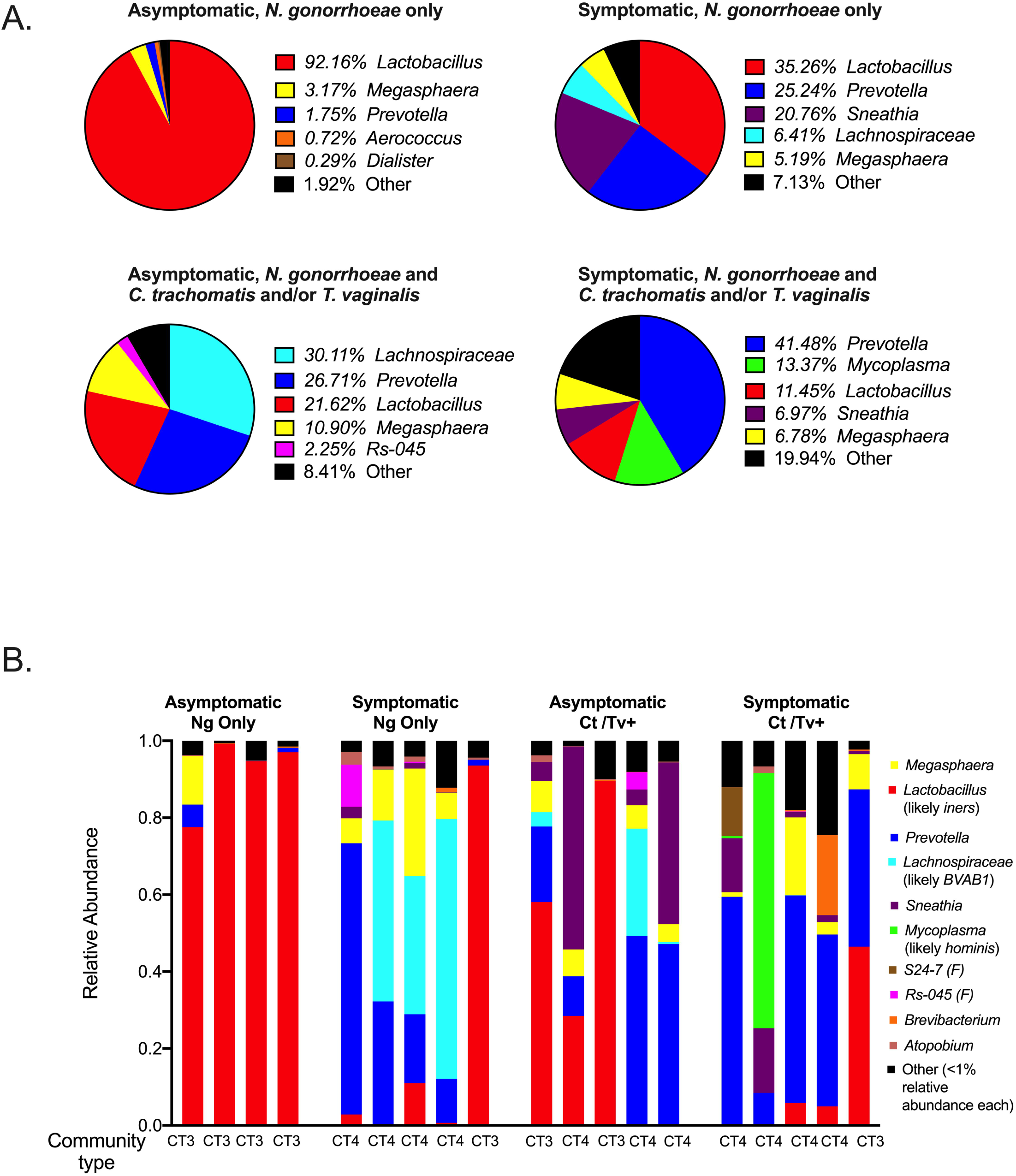
Community composition of symptomatic and asymptomatic individuals with and without *C. trachomatis* and/or *T. vaginalis* co-infection. The mean relative abundance of the top 5 most prevalent bacterial genera identified within each patient group of interest are plotted as pie charts with each pie representing a group of interest **(A)**. The relative abundances of the top ten taxa (genera or family-level taxa, as applicable) identified across all 16S rRNA sequencing reads in the dataset are shown for each of the 19 participants included in the study. These top 10 taxa comprised 299% of all reads in the entire dataset. We used BLAST and MUSCLE alignments to determine the likely species of communities dominated by *Lactobacillus, Lachnospiraceae* and *Mycoplasma* and found them to be *L. iners*, Bacterial Vaginosis-Associated Bacterium-1 (BVAB1)/”*Candidatus* Lachnocurva vaginae” and *M. hominis*, respectively. The microbial community type (CT) designated for each participant, using standard definitions in the field, is provided. **(B)**.

**Figure 5.**
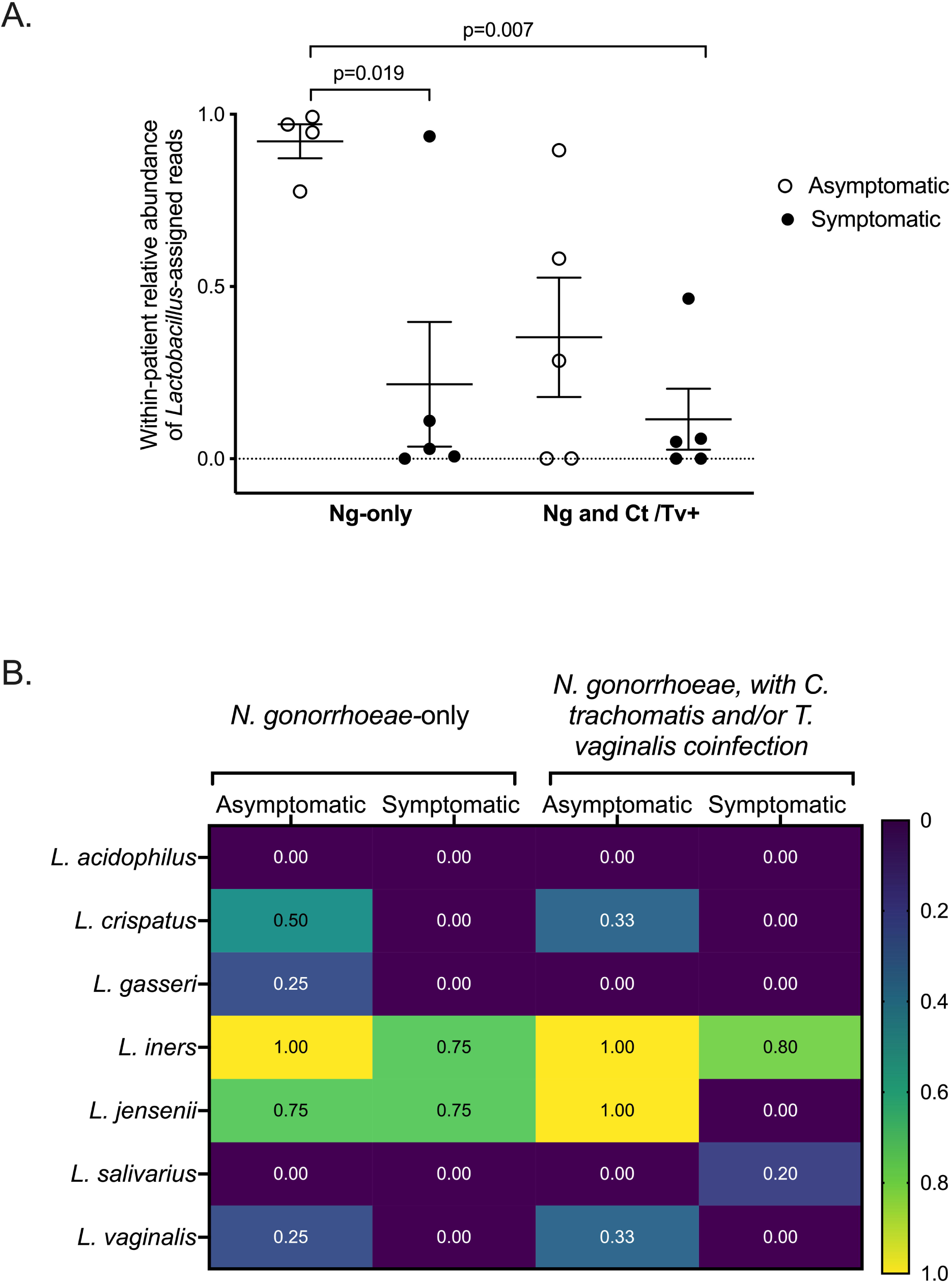
*Lactobacillus spp*. detection with respect to clinical presentation and other STI co-infection. The fraction of reads assigned to *Lactobacillus* among all sequence reads comprising an individual’s microbial community is shown **(A)**. Participants have been grouped by symptom and co-infection status. Statistical significant differences between patient groups were explored with one way ANOVA with Tukey’s correction for multiple comparisons. The presence of specific *Lactobacillus spp*. was also assessed by presence/absence commercially-available real-time PCR assay **(B)**. Participants have been grouped by symptom and co-infection status. The fraction of individuals within each group positive for each of the seven *Lactobacillus* species in our panel (*L. acidophilus, L. crispatus, L. gasseri, L. iners*,, *L. jensenii, L. salivarius, L. vaginalis*) are shown as a heatmap, with values ranging from “0” (i.e. none of the women in the group positive for that species) to “1.0” (i.e. all women in the group positive for that species).

Using BLAST and MUltiple Sequence Comparison by Log Expectation (MUSCLE) [55], we investigated the 16S sequences of each taxa that was assigned *Lactobacillus* taxonomy and identified *L. iners* as the likeliest species (Supplementary Table 1). This led us to define *Lactobacillus*-dominant samples (n=8) as community-type 3 (CT3), using standard definitions of vaginal microbial structure [21, 27, 36] (Figure 4B).

The presence of specific *Lactobacillus* species was further assessed by presence/absence real-time PCR (Figure 5B). The *Lactobacillus* species most commonly detected among all women were *L. iners, L. crispatus, L. jensenii*, and *L. gasseri* (Figure 5B). In line with our microbiome analyses and BLAST homology searches, all asymptomatic women were positive for *L. iners*, and in 85% and 75% of symptomatic women with and without co-infection, respectively. Although *L. crispatus* was not the predominant *Lactobacillus* species in any of the specimens using 16S sequencing, *L. crispatus* was detected by real-time PCR only in *N. gonorrhoeae*-infected individuals with asymptomatic presentation (Figure 5B).

### Symptomatic *N. gonorrhoeae* and STI co-infection are associated with a diverse cervicovaginal microbial community composed of bacterial vaginosis-associated bacteria

Having established that 8 of 19 samples (42.1%) were *L. iners*-dominated (CT3) and more commonly associated with asymptomatic *N. gonorrhoeae* infection, we investigated in more detail the remaining 11 samples, which were dominated by a diverse group of non-lactobacilli (*Prevotella*, a *Lachnospiraceae* genus, *Sneathia*, or *Mycoplasma*). We used BLAST and MUSCLE alignments to further characterize the composition of the non-*Lactobacillus* communities dominated by *Lachnospiraceae* and *Mycoplasma*. By applying BLAST on representative reads assigned to each genus of interest, we found that the likeliest species for *Lachnospiraceae* and *Mycoplasma* OTUs were Bacterial Vaginosis-Associated Bacterium-1 (BVAB1)/”*Candidatus* Lachnocurva vaginae” and *M. hominis*, respectively (Supplementary Table 1). Samples dominated by *Prevotella* (n=6), *Sneathia* (n=1), BVAB1 (n=3), and *M. hominis* (n=1) were classified as CT4/molecular BV. The *Mycoplasma*-dominant sample was included in the CT4 and molecular BV classifications. The CT4/molecular BV samples were more frequently found in symptomatic patients (8/10, 80% of symptomatics *vs*. 3/9, 33.3% of asymptomatics, p=0.040) (Figure 4B). Among STI co-infected individuals, 7/10 (70.0%) carried CT4 microbial communities compared to patients without co-infections (4/9, 44.4%), though this difference did not attain statistical significance (p=0.255).

The prevalence of common bacterial vaginosis-associated bacteria was also assessed by commercially-available microbial DNA real-time PCR assay (Figure 6). Of the BV-associated species included in the panel, *Gardnerella vaginalis*, was present in all samples. Other species commonly associated with BV, like *Atopobium vaginae*, certain *Prevotella spp*., and *Sneathia sanguinegens* were also highly prevalent among this cohort of women, regardless of symptoms or STI co-infection status. Asymptomatic women infected only with *N. gonorrhoeae* (n=4) carried the following species less frequently than symptomatic women infected only with *N. gonorrhoeae* (n=5): *M. hominis* (25% *vs*. 80%), *Prevotella buccalis* (25% vs. 80%), *and Ureaplasma urealyticum* (0% vs. 100%). Similarly, asymptomatic females without co-infections also carried BV-associated bacteria less frequently when compared to those with symptoms or STI co-infections (Figure 6).

**Figure 6.**
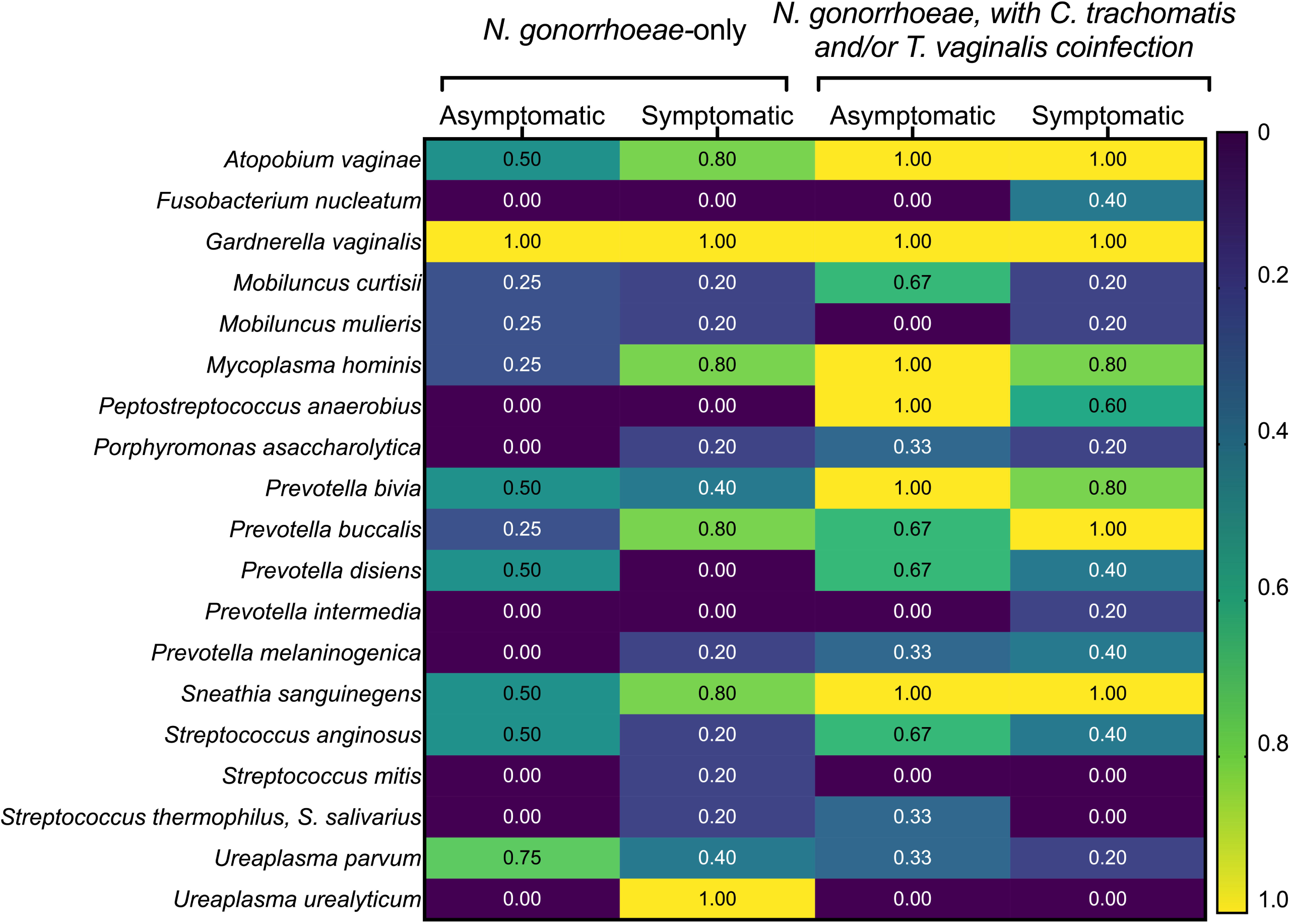
Prevalence of common bacterial vaginosis-associated bacteria detected with commercially-available microbial DNA real-time PCR assay. The presence of the indicated BV-associated bacterial species was assessed by presence/absence commercially-available real-time PCR assay. Participants have been grouped by symptom and co-infection status. The fraction of individuals within each group positive for each of the seven BV-associated bacteria in the panel are shown as a heatmap, with values ranging from “0” (i.e. none of the women in the group positive for that species) to “1.0” (i.e. all women in the group positive for that species).

**Figure 7.**
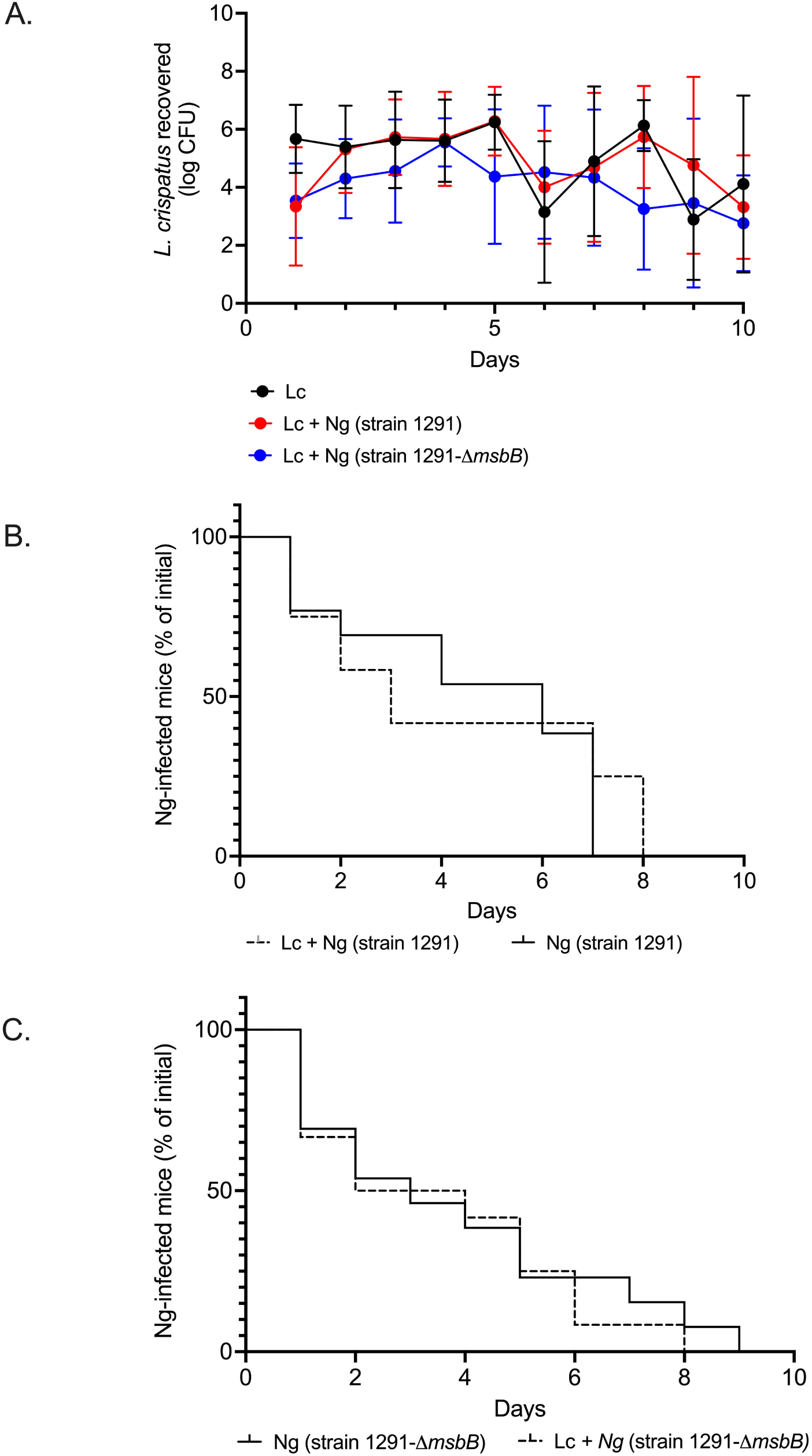
Abundance of *Lactobacillus crispatus* and *N. gonorrhoeae* during controlled gonorrhea mouse infections. Groups of 4-5 mice were pre-colonized with *L. crispatus* and then inoculated with either *N. gonorrhoeae* strain 1291 (wild-type, wt) or its isogenic strain lacking MsbB (1291-Δ*msbB*). Daily vaginal swabs were collected for 10 days and quantitative bacterial cultures for both species were performed. Results represent combined bacterial recovery findings from three independent experiments (n=3). **(A)** The average *L. crispatus* colony forming units (log_10_ CFU) recovered from daily mouse vaginal swabs are shown. The abundance of *L. crispatus* was not found to be affected by *N. gonorrhoeae* infection. **(B)**The time to clearance of *N. gonorrhoeae* strain 1291 in infected mice pre-colonized with and without *L. crispatus* was assessed with Kaplan-Meier curves. **(C)**The time to clearance of *N. gonorrhoeae* strain 1291-Δ*msbB* in infected mice pre-colonized with and without *L. crispatus* was assessed with Kaplan-Meier curves.

### A murine model does not support direct impact of *N. gonorrhoeae* on *L. crispatus* vaginal colonization

Though the microbial community in *N. gonorrhoeae* infected women was different between those with and without symptoms, it is not clear whether symptomatic infection drives reduced *Lactobacillus* abundance or whether pre-existing low *Lactobacillus* abundance or presence of BV-associated microbial communities leads to symptomatic infection. To test whether *N. gonorrhoeae* infection could affect the abundance of *Lactobacillus* present in the vagina, mice pre-colonized with *L. crispatus*, which we had found was present only in asymptomatic *N. gonorrhoeae* infected individuals, were infected with either wild type *N. gonorrhoeae* strain 1291 or an isogenic mutant strain lacking the *msbB* locus (1291Δ*msbB)*. Our group has previously shown this mutant strain induces less localized inflammation, a state that would be consistent with reduced or absent symptoms in human infection. *Lactobacillus* colonization was maintained throughout the infection in all mice, regardless of *N. gonorrhoeae* infection status. No substantial difference was observed in *Lactobacillus* burden between uninfected control mice and mice infected with either strain of *N. gonorrhoeae* (Figure 6A). To test whether the presence of *L. crispatus* differentially impacted the ability of “symptomatic” or “asymptomatic” infection inducing *N. gonorrhoeae* strains to establish and/or maintain an infection in mice, we quantified the percentage of mice infected with wild type or the Δ*msbB* strain over the course of the 10-day infection period. The presence of *L. crispatus* did not have a significant effect on the ability of the mice to clear wild-type *N. gonorrhoeae* infection (Figure 6B) or type *N. gonorrhoeae* infection with the mutant strain (Figure 6C). These findings are consistent with previous studies in mice using *N. gonorrhoeae* and *Lactobacillus* inoculation conducted on the same day [56]. Although there are caveats between mouse model and human data, our findings suggest that the presence of *L. crispatus* is not impacted by *N. gonorrhoeae* infection and that prior colonization with *L. crispatus* does not have a differential effect on *N. gonorrhoeae* infection regardless of the capacity of the *N. gonorrhoeae* to induce localized inflammation in the female mouse model of lower genital tract infection.

## Discussion

A large body of evidence links vaginal dysbiosis, such as clinical bacterial vaginosis (BV) to the risk of acquisition of several sexually-transmitted infections (STIs), including gonorrhea [17, 25, 30, 33, 57]. Despite the clear association between BV and STI acquisition risk, treatment of asymptomatic BV has not been found to reduce the incidence of *N. gonorrhoeae* or *C. trachomatis* infection incidence, raising the question whether a suboptimal vaginal environment is a modifiable biological cause of gonorrhea risk [58]. However, *Lactobacillus*-based live probiotic therapy of vaginal dysbiosis has been recently shown to reduce not only BV [16, 59], but also bacterial STI incidence [60]. To the best of our knowledge, this report is the first to examine the association between the cervicovaginal microbiota composition and symptomatic *N. gonorrhoeae* infections in women. Using 16S ribosomal RNA gene deep sequencing approaches on patient samples confirmed to be infected with *Neisseria gonorrhoeae* by APTIMA clinical testing, we show that the cervicovaginal microbiome is predictive of gonorrhea clinical presentation in women attending an STI clinic in the United States. These findings were confirmed by real-time polymerase chain reaction assays specific for several *Lactobacillus* species and BV-associated bacteria deployed in parallel on the same clinical samples.

Specimens collected from asymptomatic individuals with *N. gonorrhoeae* infection and no co-infection with *Chlamydia trachomatis* and/or *Trichomonas vaginalis* carried *Lactobacillus-*dominant microbial communities more frequently than symptomatic patients without co-infection. Notably, this *Lactobacillus* dominance was due to *L. iners* and these microbiotas were classified as community type 3 (CT3), according to established definitions in the field [21, 27, 36]. Previous studies have established that *L. iners*-dominated vaginal microbiotas compared to *L. crispatus*-dominated vaginal microbiotas may place patients more at risk of STI infection, like chlamydia or HIV [25-27]. Interestingly, none of the females in our study had cervicovaginal microbiomes dominated by *L. crispatus*, which may be consistent with a protective effect by *L. crispatus* on STI infection risk. This is supported by *in vitro* studies with clinically-isolated and lab strains of *L. crispatus* have been shown to inhibit the growth of *N. gonorrhoeae in vitro* [64], possibly through the effects of lactic acid acidification of the growth environment [65]. While *L. crispatus* produces both isomers of lactic acid, *L. iners* and human cells only make l(+) lactic acid [66, 67]. Accumulating evidence also suggests that d(−) lactic acid may impart greater protection against STI pathogens than l(+) lactic acid potentially via effects on human host cells rather than pathogen cells [68, 69]. In humans, *L. crispatus*-dominant vaginal microbiota is associated with reduced risk of acquisition of other STI, like HIV [24-27].

*N. gonorrhoeae*-infected patients who reported symptoms were found to have genital microbiomes composed of a mixture of various bacterial anaerobes, such as *Prevotella, Sneathia, Mycoplasma hominis* and Bacterial Vaginosis-Associated Bacterium-1 (BVAB1)/”*Candidatus Lachnocurva vaginae*” [70]. These women with genital microbiomes composed of anaerobes were deemed to have molecular bacterial vaginosis (BV), as defined by established classifications in the field based on diversity and relative abundance of bacterial taxa [21, 27, 36]. This included the *Mycoplasma*-dominant sample because of three main reasons that definitions of molecular BV take into account: like *Prevotella* and *Sneathia*, it can overgrow in cases of BV [71, 72], its prevalence in BV patients is three times higher compared to healthy women [73] and it is associated with severe genital mucosal inflammation [74].

A possible explanation for the association of symptoms in *N. gonorrhoeae* infection and BV-associated microbial communities relates to the known increase of inflammation and inflammatory mediators in women with BV. Several studies have shown that females with clinical BV or low *Lactobacillus* abundance and high diversity of anaerobes also harbor higher concentrations of pro-inflammatory cytokines in their genital tract [21] and higher levels of the pro-inflammatory cytokines (IL-1α, IL-1β, IL-6, IL-12 and IL-8) when compared to BV negative women [75]. Further, symptoms of abnormal vaginal discharge were also found to associate with elevated levels of IL-1β, IP-10, IL-8, and GCSF, linking inflammatory cytokines to vaginal symptoms, particularly vaginal discharge[75]. Notably, the more the vaginal microbiota shifts towards dysbiosis, the more marked the inflammation [21, 41, 42], independently of concurrent STIs, including HIV and gonorrhea [21].

We recognize that our study with clinical specimens had limitations. Because of the nature of the study design, we had no information on whether our symptomatic subjects also had clinically defined BV. However, the association between BV associated microbes and cervicovaginal community type suggests that further studies of the association of BV with symptomatic *N. gonorrhoeae* infection are needed. We lacked specimens from *N*. gonorrhoeae-negative women and the majority of our study participants identified as African American. Previous BV prevalence studies reported that compared to Caucasian women, low-*Lactobacillus* vaginal microbiotas are more common in African American and Latin women [36, 61, 62] and that up to 50% of African American women may harbor vaginal microbiotas deplete in *Lactobacillus* species [63]. Future studies on how race, the vaginal microbiota and *N. gonorrhoeae* risk intersect are needed.

Our study using human clinical specimens showed that symptomatic *vs*. asymptomatic gonorrhea presentation is correlated with having molecular BV, leading to two possible explanations. First, that the molecular BV community type compared to the *L. iners* dominated community type may have predisposed to the development of gonococcal-associated symptoms. Second, that the BV state developed after or was even caused by the establishment of *N. gonorrhoeae* infection due to the promotion of the growth of BV-associated bacteria or a loss of *Lactobacillus* species. Given the compelling evidence supporting a protective effect of *L. crispatus* on STI infection risk, we tested whether the level of *L. crispatus* colonization was impacted by either wild type or hypo-inflammatory *N. gonorrhoeae* infection in a murine model. However, no change in *L. crispatus* colonization was observed in mice infected with either *N. gonorrhoeae* strain. We also utilized this murine model to examine whether *L. crispatus*, which was only found in asymptomatic *N. gonorrhoeae*-infected humans, might reduce bacterial burden of wild-type *N. gonorrhoeae* or support the expansion of hypo-inflammatory *N. gonorrhoeae* that could theoretically be associated with asymptomatic infections. We found no impact of *L. crispatus* pre-colonization on *N. gonorrhoeae* burden from either strain. Because the murine model of *N. gonorrhoeae* infection may not fully reflect pathogenesis of *N. gonorrhoeae* infection in humans, future studies examining the microbiome longitudinally during infection are needed to provide insight into whether *N. gonorrhoeae* can influence the genital microbial composition of the host or whether pre-existing microbial composition can influence disease presentation.

Overall, our findings suggest that the cervicovaginal microbiota is a determinant, or at least a contributor, to gonorrhea clinical presentation in women. Further studies defining the relationship between genital tract microbiomes and the and pro-inflammatory immune responses in symptomatic presentation of *N. gonorrhoeae* infection are needed to elucidate whether *Lactobacilli* or BV-defining microbial communities serve as a biomarker for symptoms in *N. gonorrhoeae* infections or directly impact symptoms.

## Data Availability

Sequence reads were deposited in the Sequence Read Archive (SRA) under the accession PRJNA768436.

https://www.ncbi.nlm.nih.gov/sra/PRJNA768436

## Acknowledgments

We extend our sincere thanks to Adriane Osborne, WNP, and to the STI clinic staff and participants at the Durham County Public Health Department for making this study possible. We thank UNC Microbiome Core, and in particular Dr. M. Andrea Azcarate-Peril and Dr. Jeff Roach, for the excellent execution of the library preparation and sequencing, and subsequent analysis support.

## Ethics Statement

The studies involving human derived specimens were reviewed and approved by the University of North Carolina Institutional Review board (Studies 11-0047 and 15-2531). The research was found to meet the criteria for a waiver of informed consent for research [45 CFR 46.116(d)] and waiver of HIPAA authorization [45 CFR 164.512(i)(2)(ii)] as the study entailed research on existing specimens, posing minimal risk to participant, the waiver did not adversely affect the rights or welfare of the participants and consent/assent would have been impracticable given the loss to follow up. The mouse *N. gonorrhoeae* infection studies were reviewed and approved by the University of North Carolina at Chapel Hill Institutional Animal Care and Use Committee (IACUC ID numbers 15-217 and 18.150).

