## Supplementary figures and images for "Cervicovaginal microbiota predicts *Neisseria gonorrhoeae* clinical presentation"

### Supplementary Figure 1.tiff

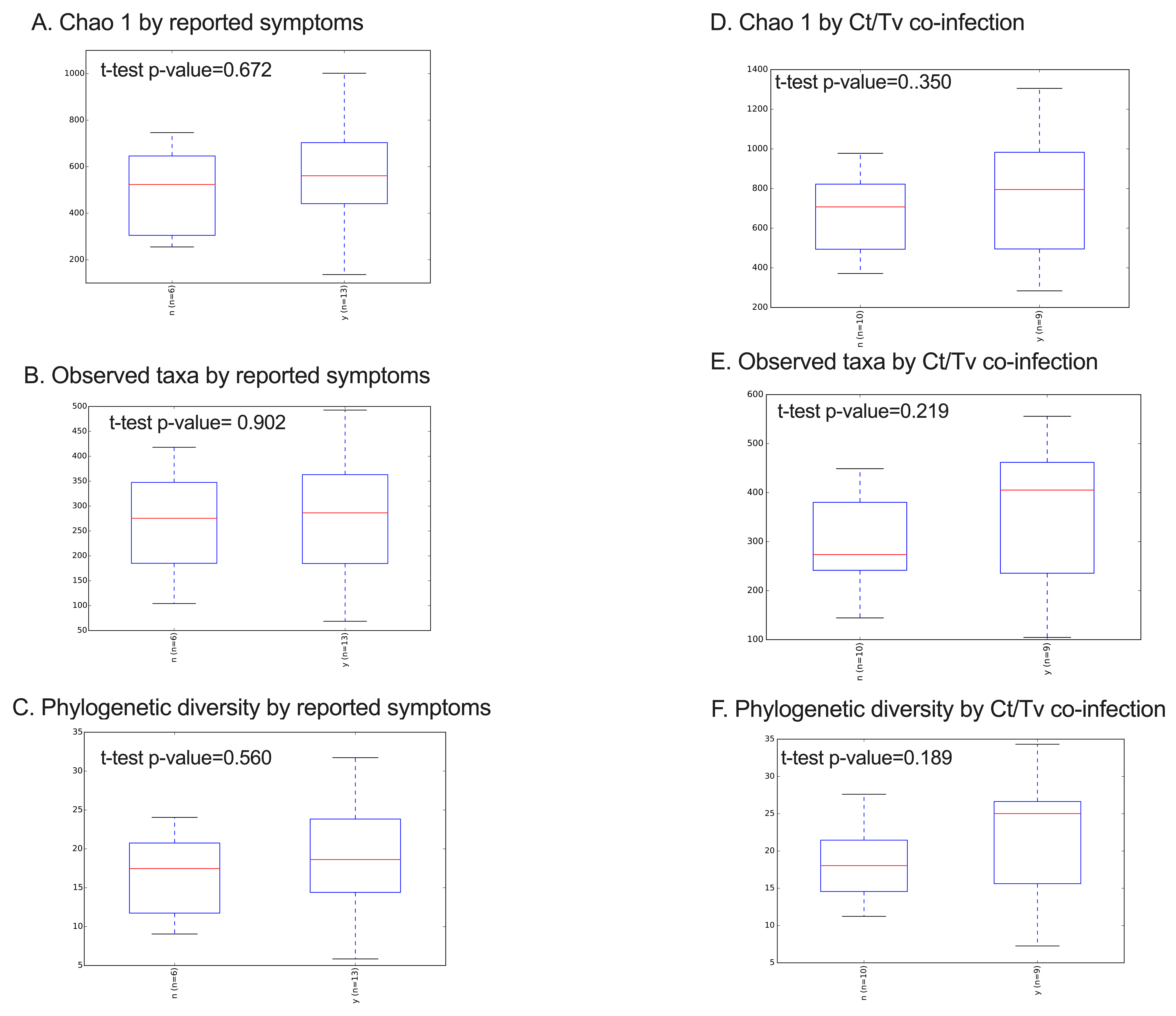
